# A systematised review of seasonal influenza case-fatality risk

**DOI:** 10.1101/2024.10.22.24315943

**Authors:** Johnny A N Filipe, Carlos K H Wong, Edwin van Leeuwen, Lucy Goodfellow, Simon R Procter, Mark Jit

**Author notes:** equal contribution (Simon R Procter, Mark Jit).

## Abstract

Case-fatality risk (CFR) is an important indicator of disease severity for influenza infection. However, estimates based on laboratory-confirmed cases (cCFR) are more-highly sensitive to features of the local health-care system and surveillance. Estimates based on diagnosed-symptomatic cases (sCFR) are more consistent across health systems but less commonly reported. We present a systematised review of sCFR for seasonal influenza. We identified 10 studies reporting sCFR, or primary data for its direct estimation, resulting in 40 location and season-specific point estimates (range 0.3-908 per 100,000 cases). The wide variation across studies and lack of studies in many world regions point to the need for standardised protocols and more data collection.

## 1. Introduction

The case-fatality risk (CFR) is a measure of disease severity and a guide to public health response to a virus outbreak or pandemic. CFR is particularly important to measure for seasonal influenza where infection severity and health care burden vary substantially among seasons, geographies and populations.

The CFR is defined as the proportion of infection-associated deaths (numerator) to infection-associated cases (denominator), but its value and interpretation depend strongly on the specific case definition. In influenza, the severity and incidence of cases follow a pyramid of categories, including (in order of increasing incidence) hospitalisations, medically-attended outpatients, symptomatic subclinical infections, and asymptomatic infections [1], [2], [3], [4]. The most useful estimator of CFR in influenza, known as infection-fatality risk (IFR), counts any infection in the community as a case, as it is more representative of the population-level health impact and free of many setting-specific factors affecting other definitions [2], [5]. However, in practice, few studies report influenza-IFR estimates due to difficulties in identifying and sampling asymptomatic or low-severity infections. Indeed, a recent literature review [6] found only one study reporting IFR for seasonal influenza, based on serological sampling [7].

Most commonly, influenza studies report CFR estimates based on laboratory-confirmed cases in hospitalised patients, and less often report estimates based on medically-attended outpatients (rarely laboratory-confirmed and occasionally supplemented with symptomatic subclinical infections [1]). These two broad estimation approaches are often termed confirmed CFR (cCFR) and symptomatic CFR (sCFR) [1], [2], [5]. However, laboratory-confirmed influenza cases (based on taking and testing a patient’s sample) are a biased indicator because outpatients are rarely tested systematically, while thresholds for hospital admission vary with local demand and resources, so there is a rationale for sCFR estimation [1], [2], [5].

Here, we present a systematised review of published population-level estimates of the sCFR, i.e. proportion of influenza-caused deaths among symptomatic seasonal-influenza cases. Studies can quantify sCFR through population or primary-care syndromic surveillance and may dependent less on the local health care system than cCFR, although they may present other forms of bias.

## 2. Methods

### Approach

We conducted a systematised review [8] of published literature reporting seasonal influenza sCFR following search and reporting procedures in the Preferred Reporting Items for Systematic Reviews and Meta-Analyses (PRISMA) statement [9]. In line with this category of review [8], we applied a pre-defined and comprehensive search strategy to identify and select literature, but did not critically evaluate the selection strategy or assess the quality of the studies included.

### Eligibility criteria

We included all full research articles reporting either a (self-defined) sCFR value for seasonal influenza or reporting symptomatic cases and mortality from the same population from which we could estimate sCFR. We allowed for a broad definition of a symptomatic case, with or without laboratory confirmation, including a surveillance record of medically diagnosed influenza-like illness (ILI) or of suspected influenza, or a report of self-diagnosed influenza symptoms (e.g. phone or online questionnaires). We allowed for data on deaths in the form of certified records or estimated excess mortality. We excluded articles that: 1) did not report on seasonal influenza; 2) did not report primary data, e.g. reviews or commentaries; 3) included hospitalised patients but not non-hospitalised symptomatic cases; 4) only reported sCFR for population subgroups (e.g. specific age or risk groups).

### Information sources

We searched published articles in PubMed and consulted relevant sources identified citing or referenced within articles we found in PubMed. All searches were conducted on 10 September 2024. We compare with IFR estimates identified in a recent literature review [6].

### Search strategy

We used the following combined search terms in the PubMed database:

(“influenza” OR “flu”) AND (“2009”[dp]:”3000”[dp]) AND “season*” AND

(((“asymptom*” OR “symptom*” OR “serolog*” OR “suspect*” OR “cases”)

AND (“infection*” OR “cases*” OR “disease” OR “burden”)

AND (“case fatality” OR “death*” OR “fatal*” OR “mortality”)

AND (“rate*” OR “risk*” OR “ratio*” OR “proportion*”))

OR

(“case fatality risk” OR “case fatality rate*” OR “case fatality ratio*”) OR (“fatality proportion” AND “case*”)))

The search was limited to articles with titles and abstracts in English language (but with any language in the main article), published between 2009 and 2024 as the influenza A(H1N1) strain prior to 2009 may have had a different CFR from the current strain.

### Selection process

The articles identified by the search were screened and evaluated by author JF to assess their eligibility against the predefined inclusion criteria. Author CW independently evaluated a 10% random sample of the articles identified and screened them to ensure consistency with JF’s evaluations. Discrepancies between the reviewers’ selected articles were resolved through discussion and joint evaluation of the full-text articles in question.

### Data collection and analysis

For each article included, a PDF-format file was downloaded from the publisher’s website and data were extracted manually from tables, main text or figures by one reviewer (JF). Data were managed and analysed in R version 4.4.1 [10]. The primary data extracted were sCFR population estimates, the underlying counts or estimates of the numbers of influenza-associated deaths (numerator) and cases (denominator), and, where provided, 95% uncertainty (confidence or credible) intervals (95%UI). If no sCFR estimates were presented, we estimated the sCFR from the numerator and denominator data; and, if interval estimates were provided for the numerator and denominator, we derived 95%UI through bootstrap sampling (R package pairwiseCI [11]). Where sCFR was reported (or calculated in this article) for over two time periods in a single study and design, we calculated their mean and derived a 95%UI assuming a Student’s t-distribution.

## 3. Results

### Studies selected

Among 1014 articles identified, we shortlisted 69 for full-text review, of which we included 9 fully meeting the eligibility criteria (**Figure 1**). We also included one additional study cited in an article found through the search (a systematic review of CFR for pandemic influenza A(H1N1) 2009 [5]). We found no other relevant systematic reviews.

**FIGURE 1.**
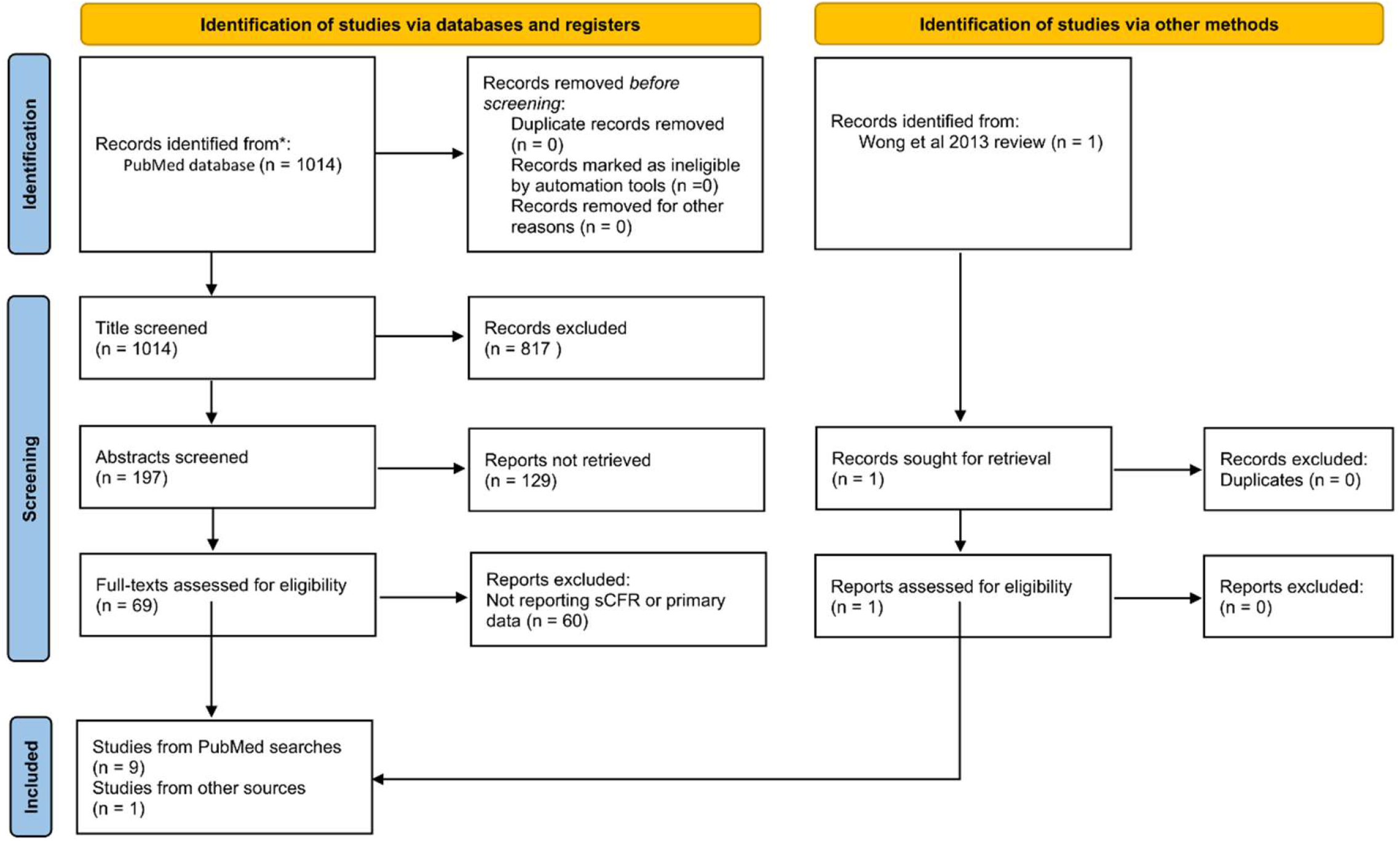
PRISMA flow diagram of the selection of studies reporting sCFR (symptomatic-case fatality risk) or primary data for its estimation.

### Study characteristics

The population characteristics and design of each study reporting sCFR for seasonal influenza, including case and death definitions, are described in **Table 1**. The studies are from 10 geographies in eastern and southern Asia, Europe, and North America; they contain a greater proportion of high-income (n=8) than middle-income (n=2) geographies, and no low-income geographies. Six in 10 studies reported case and mortality data but not sCFR estimates, and 7/10 studies did not explain or report uncertainty estimates. There is considerable heterogeneity in case definitions (e.g. ILI vs other diagnostics, symptoms in primary care vs wider community, and direct vs model-based estimates) and in death definitions (e.g. influenza-confirmed vs non-confirmed deaths, and direct vs excess mortality-based estimates). For comparison, **Table 1** includes the one study reporting IFR for seasonal influenza identified in a recent literature review [6].

**TABLE 1.**
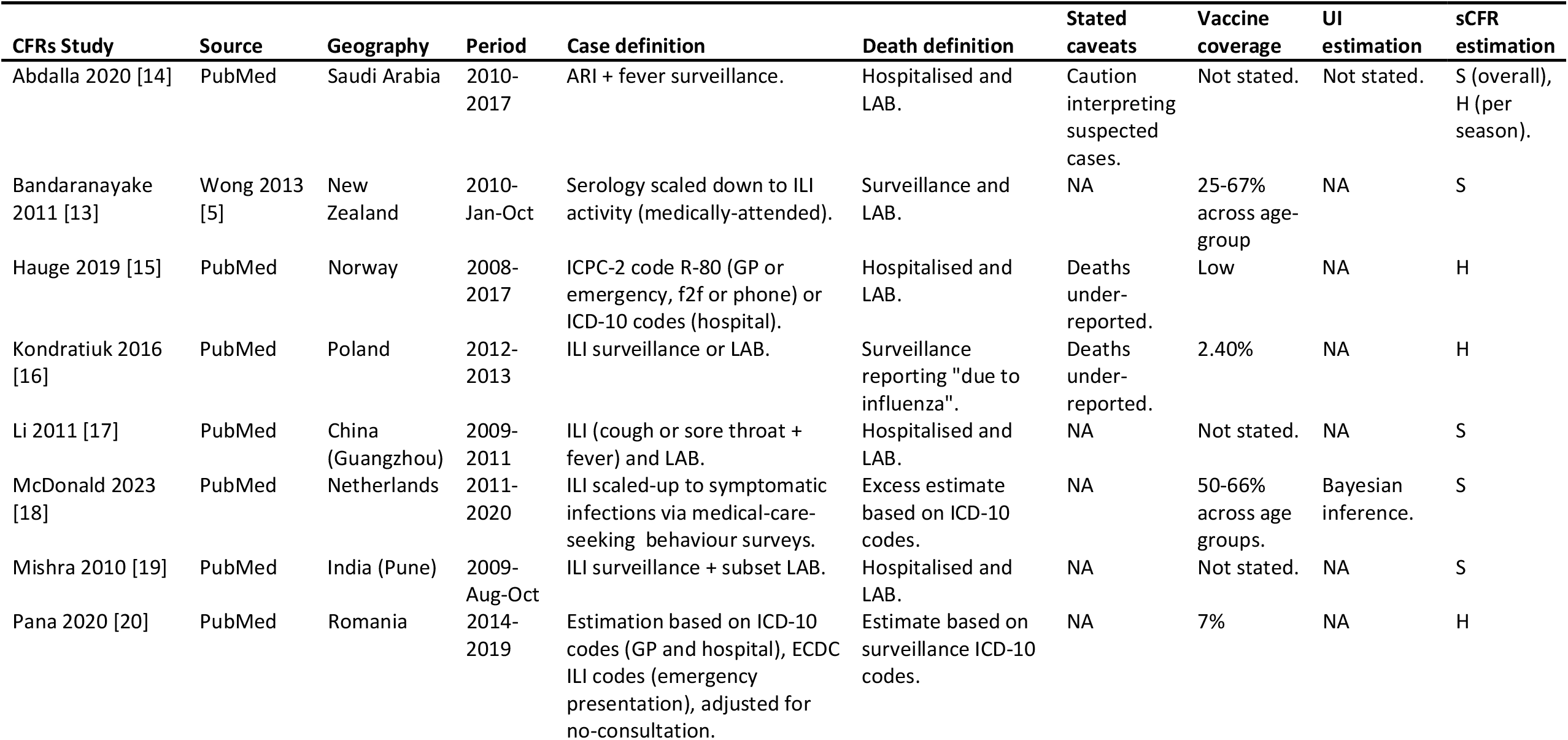
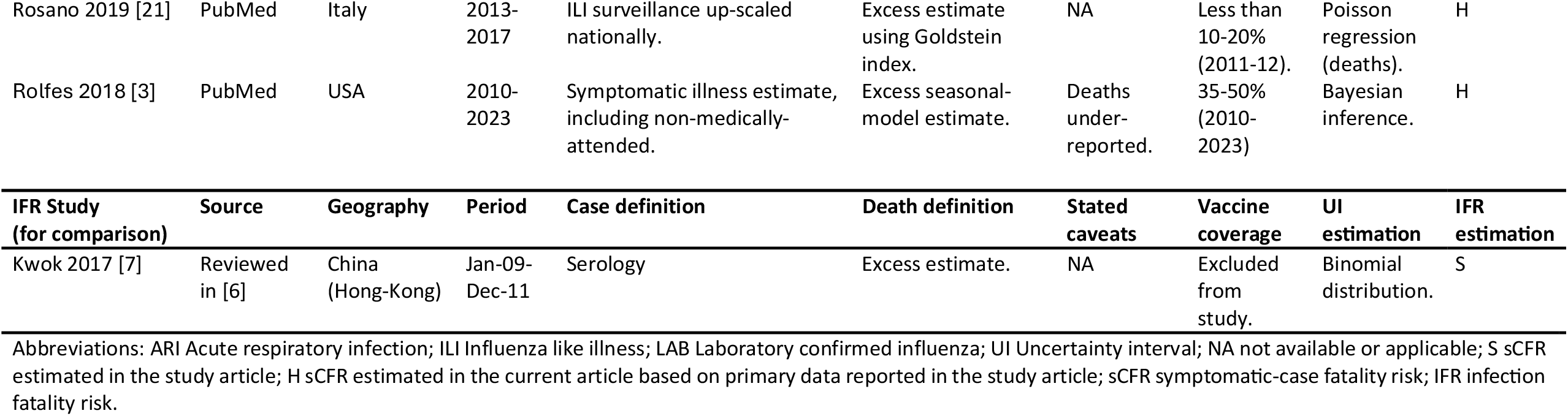
Characteristics and design of the seasonal influenza sCFR studies included in the review, and, for comparison, of the seasonal influenza IFR study identified in a previous review.

### sCFR values for individual studies

A joint plot and table of sCFR estimates from the 10 studies included is shown (**Figure 2**). We did not display uncertainty intervals as the stated uncertainty ranges in each study are not directly comparable (they were estimated using different methods (**Table 1**)). There are 40 sCFR point estimates across locations and seasons (range 0.3-907.7, IQR 70.5-191.3 per 100,000 cases). In comparison, a systematic review for pandemic influenza A(H1N1) 2009 [1] included 25 studies and 30 sCFR point estimates (range 0-1200, IQR 6.8-29.0; or, removing outliers 1200 and 440 (Mexico), range 0-100, IQR 6.4-24.5 per 100,000 cases; see Supporting Information); the seasonal estimates are overall larger than those for the 2009 pandemic. Four studies (Italy, Norway, Saudi Arabia and USA) have sCFR estimates from multiple (more than two) seasons and two studies (China (Guangzhou) and Poland) have estimates from two seasons. Geographies with multiple estimates show substantial seasonal variation within location, wider than that across most geographies. In addition, the sCFR is expected (based on previous literature [5]) to be about one order of magnitude greater than the IFR, which overall is the case when comparing with the single seasonal-influenza IFR estimate [7] identified in [6].

**FIGURE 2.**
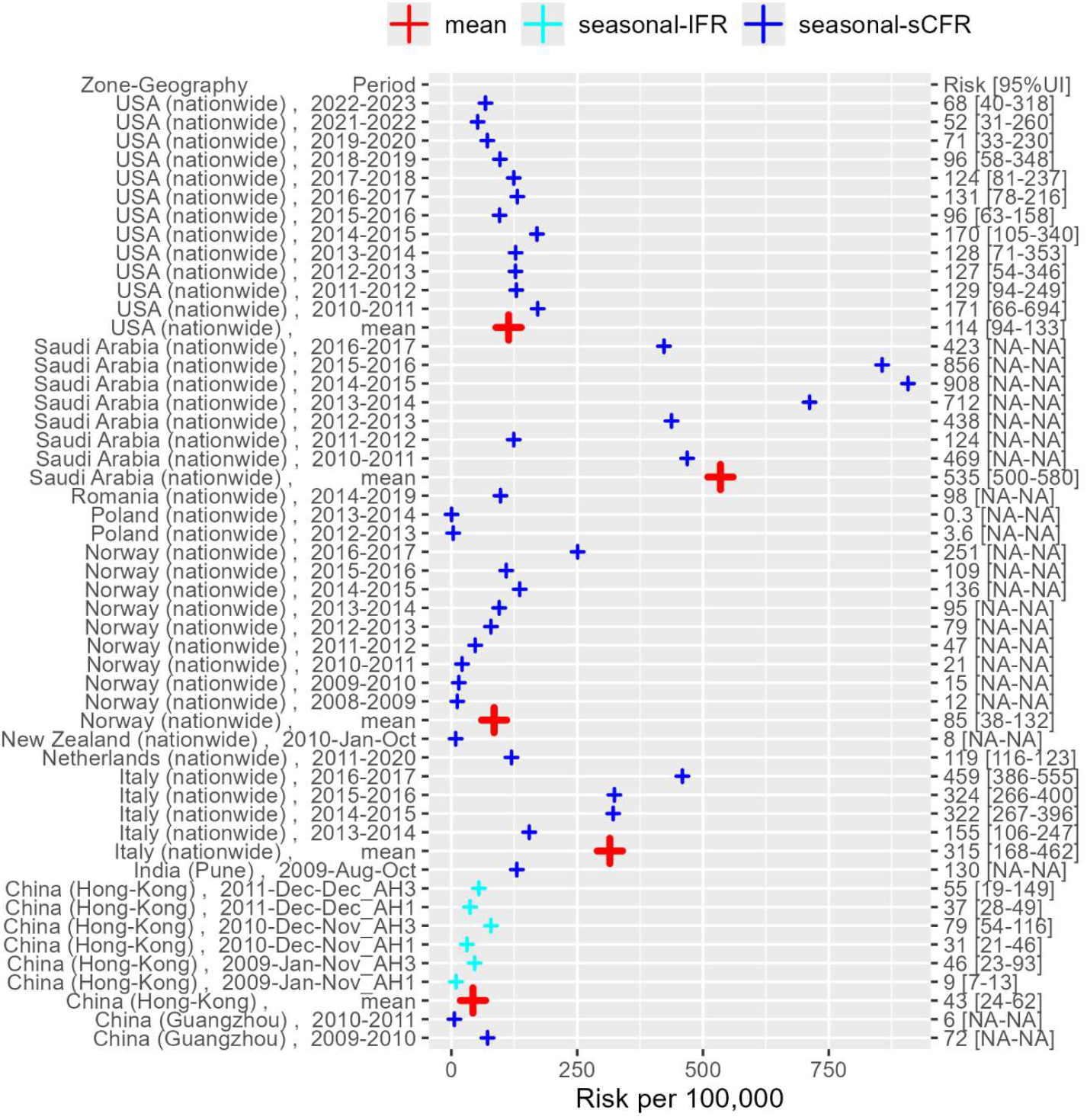
Table and plot of the sCFR (symptomatic-case fatality risk) estimates from the studies included in the review. For comparison, an IFR estimate for Hong-Kong identified in a recent review in [6] is included. Each mean is over the periods in the same geography and study (if more than two periods). Uncertainty intervals are stated but not plotted as they are not comparable (see Results and Table 1). Abbreviations: AH1: influenza A(H1N1), AH3: influenza A(H3N1); UI: uncertainty interval; NA: not applicable.

## 4. Discussion

Only 10 studies were identified reporting seasonal influenza sCFR. These studies are predominantly based in high-income geographies, half of which are European. The sCFR point estimates are overall larger than those for pandemic influenza A(H1N1) 2009 [5], in agreement with findings that the 2009 pandemic was milder than seasonal influenza in subsequent seasons [1], [12]. On the other hand, the sCFR estimates were considerably larger than the only seasonal-influenza IFR estimate previously identified, consistently with expectation [2], [5], although the geography on which the IFR study was based (Hong Kong) has no sCFR estimate to compare against.

Studies with sCFR estimates from multiple seasons indicate that the range across seasons can be wider than the range across geographies, highlighting the importance of measuring sCFR seasonally as well as locally. Such seasonal variation may be due to change in season start and duration, strain severity, vaccine coverage and vaccine strain matching (where a vaccination programme exists) [13]. Some of the sCFR variation between studies may also be due to heterogeneity in surveillance systems, case and death definitions, population characteristics, climate-specific seasonal pattern, and vaccination coverage (if any) [2], [4], [5]. More specifically, the symptomatic-case estimates (denominator) may have over-ascertainment bias (causing sCFR under-ascertainment) through the inclusion of non-influenza ILI symptoms if not all cases were laboratory confirmed, or under-ascertainment bias through the exclusion of subclinical symptomatic infections if primary-care cases were included but non-medically-attended cases were not (their relative proportions can be estimated e.g. via surveys). The death estimates (numerator in sCFR) may also have under-ascertainment bias through underreporting or misdiagnosis, or over-ascertainment if excess rather than confirmed mortality was used. The potential biases are not necessarily discernible by directly comparing sCFR estimates among the current studies. An ideal study design would include comprehensive primary-care and outpatient surveillance, laboratory confirmation of cases and deaths, and adjustment for non-medically-attended symptomatic cases [5].

In conclusion, sCFR estimates for seasonal influenza show consistency with previous IFR and pandemic-influenza studies [5], [6] and are more informative of their range when inclusive of multiple seasons. However, large study variation, methodological limitations (e.g. unreported sCFR point or uncertainty estimates), and the lack of studies in many world regions point to the need for more studies and more consistent data collection.

## Author Contributions

**Johnny Filipe:** Conceptualization; methodology; investigation; software; data curation; formal analysis; visualization; writing – original draft; writing – review and editing. **Carlos Wong:** Conceptualization; methodology; investigation; writing – review and editing. **Edwin van Leeuwen:** Methodology; writing – review and editing. **Lucy Goodfellow:** Visualization; writing – review and editing. **Simon Procter:** Conceptualization; methodology; supervision; writing – review and editing. **Mark Jit:** Conceptualization; methodology; supervision; writing – review and editing.

## Acknowledgements

We thank Rosalind Eggo (London School of Hygiene and Tropical Medicine) and Joseph Wu (University of Hong Kong) for their involvement in obtaining funding for this work, and for their input in the conceptualisation of the wider project that inspired the current work.

LG, SP and MJ were supported by the Task Force for Global Health in collaboration with Partnership for Influenza Vaccine Introduction (PIVI). JF, CW and MJ were supported by AIR@InnoHK administered by the Innovation and Technology Commission of the Government of the Hong Kong Special Administrative Region, China, as part of the Laboratory of Data Discovery for Health (D24H). EvL was supported by the National Institute for Health Research (NIHR) Health Protection Research Unit (HPRU) in Modelling and Health Economics, a partnership between UKHSA, Imperial College London, and LSHTM (grant number NIHR200908). The views expressed are those of the authors and not necessarily those of the UK Department of Health and Social Care (DHSC), NIHR, or UKHSA.

## Data availability statement

The sCFR estimates used in this review are shown in tables and their primary data sources are cited. In addition, the primary data extracted and the codes for their analysis and for outputting tables and figures are available in the repository https://github.com/JAN-Filipe/scfr-influenza

## Supporting Information

Additional supporting information can be found online in the Supporting Information section.

## References

[1] A. M. Presanis et al., “The Severity of Pandemic H1N1 Influenza in the United States, from April to July 2009: A Bayesian Analysis,” PLOS Med., vol. 6, no. 12, p. e1000207, Dec. 2009, doi: 10.1371/journal.pmed.1000207.

[2] H. Nishiura, “The virulence of pandemic influenza A (H1N1) 2009: an epidemiological perspective on the case–fatality ratio,” Expert Rev. Respir. Med., vol. 4, no. 3, pp. 329–338, Jun. 2010, doi: 10.1586/ers.10.24.

[3] M. A. Rolfes et al., “Annual estimates of the burden of seasonal influenza in the United States: A tool for strengthening influenza surveillance and preparedness,” Influenza Other Respir. Viruses, vol. 12, no. 1, pp. 132–137, Jan. 2018, doi: 10.1111/irv.12486.

[4] C. Pelat et al., “Optimizing the precision of case fatality ratio estimates under the surveillance pyramid approach.,” Am. J. Epidemiol., vol. 180, no. 10, pp. 1036–1046, Nov. 2014, doi: 10.1093/aje/kwu213.

[5] J. Y. Wong, H. Kelly, D. K. M. Ip, J. T. Wu, G. M. Leung, and B. J. Cowling, “Case Fatality Risk of Influenza A (H1N1pdm09): A Systematic Review,” Epidemiology, vol. 24, no. 6, 2013, [Online]. Available: https://journals.lww.com/epidem/fulltext/2013/11000/case_fatality_risk_of_influenza_ah1n1pdm09a.6.aspx

[6] L. Goodfellow et al., “The potential global impact and cost-effectiveness of next-generation influenza vaccines: a modelling analysis,” medRxiv, p. 2024.09.19.24313950, Jan. 2024, doi: 10.1101/2024.09.19.24313950.

[7] K. O. Kwok et al., “Relative incidence and individual-level severity of seasonal influenza A H3N2 compared with 2009 pandemic H1N1.,” BMC Infect. Dis., vol. 17, no. 1, p. 337, May 2017, doi: 10.1186/s12879-017-2432-7.

[8] M. J. Grant and A. Booth, “A typology of reviews: an analysis of 14 review types and associated methodologies,” Health Inf. Libr. J., vol. 26, no. 2, pp. 91–108, Jun. 2009, doi: 10.1111/j.1471-1842.2009.00848.x.

[9] M. J. Page et al., “The PRISMA 2020 statement: an updated guideline for reporting systematic reviews,” BMJ, vol. 372, p. n71, Mar. 2021, doi: 10.1136/bmj.n71.

[10] R Core Team, R: A Language and Environment for Statistical Computing. (2024). R Foundation for Statistical Computing, Vienna, Austria. [Online]. Available: https://www.R-project.org/

[11] F. Schaarschmidt, “pairwiseCI: Confidence Intervals for Two Sample Comparisons.” 2024. doi: 10.32614/CRAN.package.pairwiseCI.

[12] M. Athanasiou et al., “Influenza surveillance during the post-pandemic influenza 2010/11 season in Greece, 04 October 2010 to 22 May 2011.,” Euro Surveill. Bull. Eur. Sur Mal. Transm. Eur. Commun. Dis. Bull., vol. 16, no. 44, p. 20004, Nov. 2011.

[13] D. Bandaranayake et al., “The second wave of 2009 pandemic influenza A(H1N1) in New Zealand, January–October 2010,” Eurosurveillance, vol. 16, no. 6. p. 19788, 2011.

[14] O. Abdalla et al., “Hospital-based surveillance of influenza A(H1N1)pdm09 virus in Saudi Arabia, 2010-2016.,” Ann. Saudi Med., vol. 40, no. 1, pp. 1–6, Feb. 2020, doi: 10.5144/0256-4947.2020.1.

[15] S. H. Hauge, I. J. Bakken, B. F. de Blasio, and S. E. Håberg, “Burden of medically attended influenza in Norway 2008-2017,” Influenza Other Respir. Viruses, vol. 13, no. 3, pp. 240–247, May 2019, doi: 10.1111/irv.12627.

[16] K. Kondratiuk et al., “Influenza in Poland in 2013 and 2013/2014 epidemic season.,” Przegl. Epidemiol., vol. 70, no. 3, pp. 407–419, 2016.

[17] T. Li et al., “A two-year surveillance of 2009 pandemic influenza A (H1N1) in Guangzhou, China: from pandemic to seasonal influenza?,” PloS One, vol. 6, no. 11, p. e28027, 2011, doi: 10.1371/journal.pone.0028027.

[18] S. A. McDonald et al., “Inference of age-dependent case-fatality ratios for seasonal influenza virus subtypes A(H3N2) and A(H1N1)pdm09 and B lineages using data from the Netherlands,” Influenza Other Respir. Viruses, vol. 17, no. 6, p. e13146, Jun. 2023, doi: 10.1111/irv.13146.

[19] A. C. Mishra, M. S. Chadha, M. L. Choudhary, and V. A. Potdar, “Pandemic influenza (H1N1) 2009 is associated with severe disease in India.,” PloS One, vol. 5, no. 5, p. e10540, May 2010, doi: 10.1371/journal.pone.0010540.

[20] A. Pană, A. Pistol, A. Streinu-Cercel, and B.-V. Ileanu, “Burden of influenza in Romania. A retrospective analysis of 2014/15 - 2018/19 seasons in Romania.,” Germs, vol. 10, no. 4, pp. 201–209, Sep. 2020, doi: 10.18683/germs.2020.1206.

[21] A. Rosano et al., “Investigating the impact of influenza on excess mortality in all ages in Italy during recent seasons (2013/14–2016/17 seasons),” Int. J. Infect. Dis., vol. 88, pp. 127–134, Nov. 2019, doi: 10.1016/j.ijid.2019.08.003.

