## Supplementary Table S1 and Figure S1 for "A systematised review of seasonal influenza case-fatality risk"

* equal contribution

The following data are estimates of symptom-based case fatality-risk extracted from a previous literature review on CFR for pandemic influenza A(H1N1) 2009 [1].

**Table S1 –** Studies reporting sCFR for pandemic influenza A(H1N1) 2009 in Wong et al 2013. sCFR range 0-1200 and IQR 6.8-29.0 per 100,000 cases (range 0-100 and IQR 6.4-24.5 if removing outliers 1200 and 440 (Mexico)).

| Study | Geography | Period | sCFR/100,000 | 95%UI |
|---|---|---|---|---|
| Abdalla 2010 [2] | USA | Apr-09-Apr-10 | 20.4 | NA |
| Baker 2009 [3] | New Zealand | Jun-Aug-2009 | 5 | 3-11 |
| Bandaranayake 2010 [4] | New Zealand | Apr-Sep-2009 | 8.2 | NA |
| Brooks-Pollock 2011 [5] | UK (England) | Jul-Nov-2009 | 17 | NA |
| Chowell 2011 [6] | Mexico | Apr-Dec-2009 | 1200 | 1100-1200 |
| Cutter 2010 [7] | Mexico | Jun-Oct-2009 | 6.7 | NA |
| Dawood 2010 [8] | Australia | Jun-Aug-2009 | 9.4 | 7.1-13.2 |
| Donaldson 2009 [9] | UK (England) | Jun-Nov-2009 | 26 | 11-66 |
| Doshi 2012 [10] | USA (Atlanta) | Aug-Sep-2009 | 24 | NA |
| Echevarria-Zuno [11] | Mexico | Apr-Jul-2009 | 100 | NA |
| Flahault 2009 [12] | New Caledonia | Aug-2009 | 10 | NA |
| | Mauritius | Aug-2009 | 10 | NA |
| Fraser 2009 [13] | Mexico | Mar-Apr-2009 | 440 | 370-520 |
| Godoy 2011 [14] | Spain | Jun-09-May-10 | 30 | 10-40 |
| Hadler 2010 [15] | USA | May-Jun-2009 | 5.4 | 4.7-6.5 |
| Kamigaki 2009 [16] | Japan | Jul-Dec-2009 | 0.67 | NA |
| Kim 2011 [17] | South Korea | Aug-Nov-2009 | 16 | NA |
| Larrieu 2011 [18] | Martinique | Aug-09-Jan-10 | 5 | NA |
| | Guadalupe | Aug-09-Jan-10 | 31 | NA |
| | French Guiana | Aug-09-Jan-10 | 17 | NA |
| | St Martin | Aug-09-Jan-10 | 0 | NA |
| Mendez 2011 [19] | Spain | May-09-Mar-10 | 43 | 38-48 |
| Nishiura 2010 [20] | Japan | Jul-09-Jan-10 | 0.94 | 0.8–1.08 |
| Pebody 2010 [21] | UK | Apr-09-Mar-10 | 40 | 20-100 |
| Presanis 2009 [22] | USA | Apr-Jul-2009 | 48 | 26-96 |

| | | | | |
|---|---|---|---|---|
| | USA (SR-ILI) | Apr-Jul-2009 | 7 | 5-9 |
| Presanis 2011 [23] | UK | Jun-Aug-2009 | 15 | 10-22 |
| Renault 2011 [24] | Reunion Island | Jul-09-Oct-09 | 7 | NA |
| Sypsa 2011 [25] | Greece | Aug-09-Feb-10 | 17.5 | 14.6-20.8 |
| Wilson 2009 [26] | High income | Apr-Jun-2009 | 2.5 | NA |

Abbreviations: sCFR symptomatic-case fatality-risk; UI uncertainty interval; NA not available; SR-ILI self-reported ILI from surveys; IQR inter quartile range.

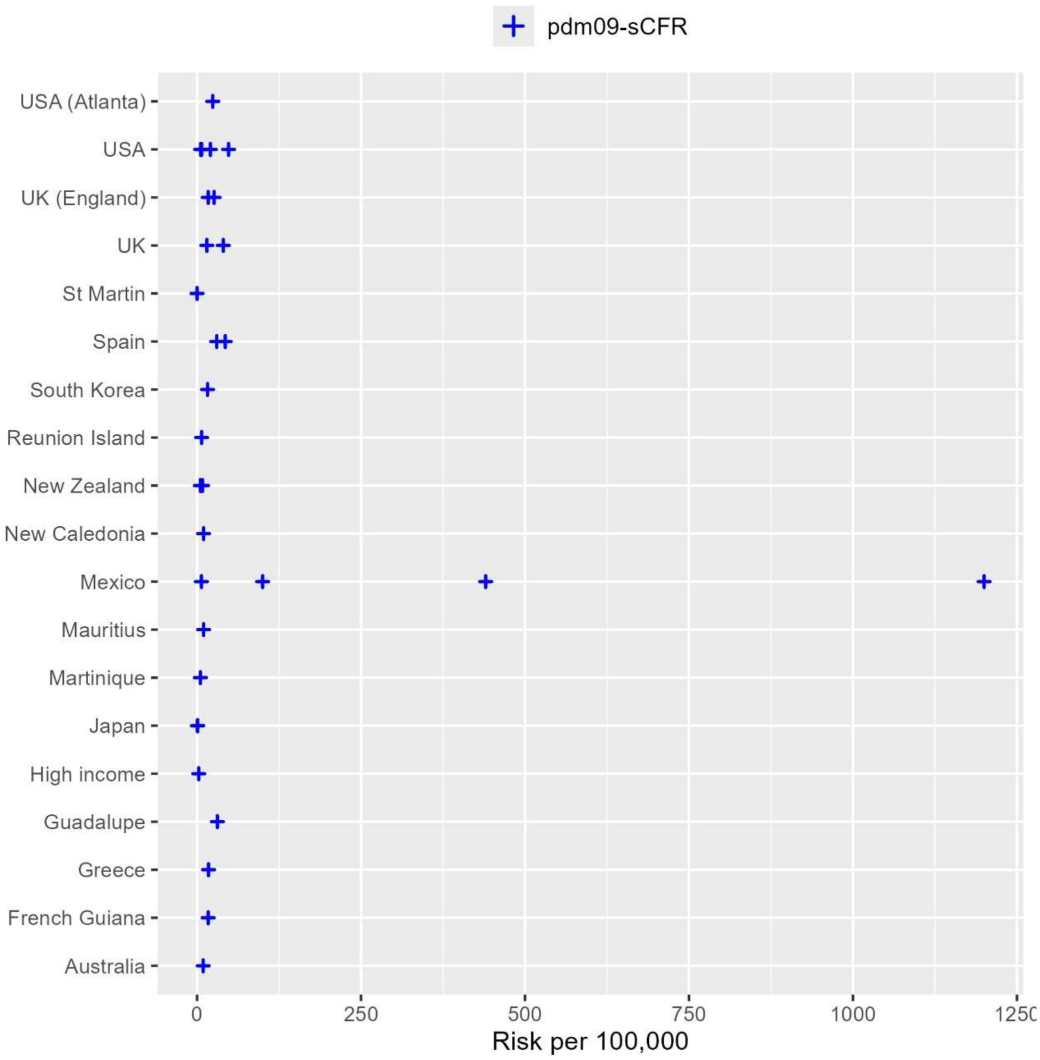


**Figure S1 –** Plot of sCFR (symptomatic-case fatality risk) point estimates reported in Wong et al 2013 (Table S1 shows the values of the point and 95%UI estimates).

### References


[1] J. Y. Wong, H. Kelly, D. K. M. Ip, J. T. Wu, G. M. Leung, and B. J. Cowling, "Case Fatality Risk of Influenza A (H1N1pdm09): A Systematic Review," *Epidemiology*, vol. 24, no. 6, 2013, [Online]. Available:

https://journals.lww.com/epidem/fulltext/2013/11000/case_fatality_risk_of_influenza_a__h1n1pdm09___a.6.aspx

[2] E. Abdalla *et al.*, “Epidemiology of influenza A 2009 H1N1 virus pandemic in the U.S.,” *J. Health Care Poor Underserved*, vol. 22, no. 4 Suppl, pp. 39–60, 2011, doi: 10.1353/hpu.2011.0163.

[3] M. G. Baker *et al.*, “Pandemic influenza A(H1N1)v in New Zealand: the experience from April to August 2009.,” *Euro Surveill. Bull. Eur. Sur Mal. Transm. Eur. Commun. Dis. Bull.*, vol. 14, no. 34, p. 19319, Aug. 2009, doi: 10.2807/ese.14.34.19319-en.

[4] D. Bandaranayake *et al.*, “Risk factors and immunity in a nationally representative population following the 2009 influenza A(H1N1) pandemic.,” *PloS One*, vol. 5, no. 10, p. e13211, Oct. 2010, doi: 10.1371/journal.pone.0013211.

[5] E. Brooks-Pollock, N. Tilston, W. J. Edmunds, and K. T. Eames, “Using an online survey of healthcare-seeking behaviour to estimate the magnitude and severity of the 2009 H1N1v influenza epidemic in England,” *BMC Infect. Dis.*, vol. 11, no. 1, p. 68, Mar. 2011, doi: 10.1186/1471-2334-11-68.

[6] G. Chowell *et al.*, “Characterizing the epidemiology of the 2009 influenza A/H1N1 pandemic in Mexico.,” *PLoS Med.*, vol. 8, no. 5, p. e1000436, May 2011, doi: 10.1371/journal.pmed.1000436.

[7] J. L. Cutter, L. W. Ang, F. Y. L. Lai, H. Subramony, S. Ma, and L. James, “Outbreak of pandemic influenza A (H1N1-2009) in Singapore, May to September 2009.,” *Ann. Acad. Med. Singapore*, vol. 39, no. 4, pp. 273–210, Apr. 2010.

[8] F. S. Dawood, K. G. Hope, D. N. Durrheim, R. Givney, A. M. Fry, and C. B. Dalton, “Estimating the disease burden of pandemic (H1N1) 2009 virus infection in Hunter New England, Northern New South Wales, Australia, 2009.,” *PloS One*, vol. 5, no. 3, p. e9880, Mar. 2010, doi: 10.1371/journal.pone.0009880.

[9] L. J. Donaldson *et al.*, “Mortality from pandemic A/H1N1 2009 influenza in England: public health surveillance study.,” *BMJ*, vol. 339, p. b5213, Dec. 2009, doi: 10.1136/bmj.b5213.

[10] S. S. Doshi *et al.*, “The Burden and Severity of Illness Due to 2009 Pandemic Influenza A (H1N1) in a Large US City During the Late Summer and Early Fall of 2009,” *Am. J. Epidemiol.*, vol. 176, no. 6, pp. 519–526, Sep. 2012, doi: 10.1093/aje/kws137.

[11] S. Echevarría-Zuno *et al.*, “Infection and death from influenza A H1N1 virus in Mexico: a retrospective analysis.,” *Lancet Lond. Engl.*, vol. 374, no. 9707, pp. 2072–2079, Dec. 2009, doi: 10.1016/S0140-6736(09)61638-X.

[12] A. Flahault, “First estimation of direct H1N1pdm virulence: From reported non consolidated data from Mauritius and New Caledonia.,” *PLoS Curr.*, vol. 1, p. RRN1010, Aug. 2009, doi: 10.1371/currents.rrn1010.

[13] C. Fraser *et al.*, “Pandemic potential of a strain of influenza A (H1N1): early findings.,” *Science*, vol. 324, no. 5934, pp. 1557–1561, Jun. 2009, doi: 10.1126/science.1176062.

[14] P. Godoy *et al.*, “[Surveillance of the pandemic influenza (H1N1) 2009 in Catalonia: results and implications].,” *Rev. Esp. Salud Publica*, vol. 85, no. 1, pp. 37–45, Feb. 2011, doi: 10.1590/S1135-57272011000100005.

[15] J. L. Hadler *et al.*, “Case fatality rates based on population estimates of influenza-like illness due to novel H1N1 influenza: New York City, May-June 2009.,” *PloS One*, vol. 5, no. 7, p. e11677, Jul. 2010, doi: 10.1371/journal.pone.0011677.

[16] T. Kamigaki and H. Oshitani, “Epidemiological characteristics and low case fatality rate of pandemic (H1N1) 2009 in Japan.,” *PLoS Curr.*, vol. 1, p. RRN1139, Dec. 2009, doi: 10.1371/currents.RRN1139.

[17] H. S. Kim *et al.*, “Fatal cases of 2009 pandemic influenza A (H1N1) in Korea.,” *J. Korean Med. Sci.*, vol. 26, no. 1, pp. 22–27, Jan. 2011, doi: 10.3346/jkms.2011.26.1.22.

[18] S. Larrieu *et al.*, “[Epidemic of influenza A(H1N1) 2009 in the French overseas territories of the Americas: epidemiological surveillance set up and main results, April 2009-January 2010].,” *Bull. Soc. Pathol. Exot. 1990*, vol. 104, no. 2, pp. 119–124, May 2011, doi: 10.1007/s13149-010-0111-7.

[19] L. Simón Méndez, S. de Mateo Ontañón, A. Larrauri Cámara, S. Jiménez-Jorge, J. Vaqué Rafart, and S. Pérez Hoyos, “[Transmissibility and severity of the pandemic influenza A (H1N1) 2009 virus in Spain].,” *Gac. Sanit.*, vol. 25, no. 4, pp. 296–302, Aug. 2011, doi: 10.1016/j.gaceta.2011.02.008.

[20] H. Nishiura, “The virulence of pandemic influenza A (H1N1) 2009: an epidemiological perspective on the case–fatality ratio,” *Expert Rev. Respir. Med.*, vol. 4, no. 3, pp. 329–338, Jun. 2010, doi: 10.1586/ers.10.24.

[21] R. G. Pebody *et al.*, “Pandemic Influenza A (H1N1) 2009 and mortality in the United Kingdom: risk factors for death, April 2009 to March 2010.,” *Euro Surveill. Bull. Eur. Sur Mal. Transm. Eur. Commun. Dis. Bull.*, vol. 15, no. 20, p. 19571, May 2010.

[22] A. M. Presanis *et al.*, “The Severity of Pandemic H1N1 Influenza in the United States, from April to July 2009: A Bayesian Analysis,” *PLOS Med.*, vol. 6, no. 12, p. e1000207, Dec. 2009, doi: 10.1371/journal.pmed.1000207.

[23] A. M. Presanis *et al.*, “Changes in severity of 2009 pandemic A/H1N1 influenza in England: a Bayesian evidence synthesis,” *BMJ*, vol. 343, p. d5408, Sep. 2011, doi: 10.1136/bmj.d5408.

[24] P. Renault *et al.*, “[Epidemic of influenza A(H1N1) 2009 in Reunion Island: epidemiological data].,” *Bull. Soc. Pathol. Exot. 1990*, vol. 104, no. 2, pp. 108–113, May 2011, doi: 10.1007/s13149-010-0113-5.

[25] V. Sypsa *et al.*, “Estimating the disease burden of 2009 pandemic influenza A(H1N1) from surveillance and household surveys in Greece.,” *PloS One*, vol. 6, no. 6, p. e20593, 2011, doi: 10.1371/journal.pone.0020593.

[26] N. Wilson and M. G. Baker, “The emerging influenza pandemic: estimating the case fatality ratio,” *Eurosurveillance*, vol. 14, no. 26. p. 19255, 2009.
